# Association of Human Mobility and Weather Conditions with Dengue Mosquito Abundance during the COVID-19 Pandemic in Hong Kong

**DOI:** 10.1101/2024.04.17.24306004

**Authors:** Yufan Zheng, Keqi Yue, Eric W. M. Wong, Hsiang-Yu Yuan

## Abstract

**Background:** While *Aedes* mosquitoes, the Dengue vectors, were expected to expand their spread due to international travel and climate change, the effects of human mobility and low rainfall conditions on them are largely unknown. We aimed to assess these influences during the COVID-19 pandemic in Hong Kong, characterized by varying levels of human mobility.

**Methods:** Google’s human mobility indices (including residential, parks and workplaces) and weather conditions (total rainfall and mean temperature) together with *Aedes albopictus* abundance and extensiveness monitored using Gravidtrap were obtained between April 2020 and August 2022. Distributed lag non-linear models with mixed-effects models were used to explore their influence in three areas in Hong Kong.

**Findings:** The relative risk (RR) of mosquito abundance was associated with low rainfall (<50 mm) after 4.5 months, with a maximum of 1.73, compared with 300 mm. Heavy rainfall (>500 mm) within 3 months was also associated with a peak of RR at 1.41. Warm conditions (21-30°C; compared with 20°C) were associated with a higher RR of 1.47 after half a month. Residential mobility was negatively associated with mosquito abundance. The model projected that if residential mobility in the year 2022 was reduced to the level before the COVID-19 pandemic, the mosquito abundance would increase by an average of 80.49% compared to the actual observation.

**Significance:** Both the human mobility and the lag effect of meteorological factors can be critical for the prediction of vector dynamics, and stay-at-home policy may be useful for its control in certain regions.

**AUTHOR SUMMARY:** Previous studies have demonstrated that both meteorological factors and human mobility were linked to the risk of Dengue transmission, with rainfall potentially exerting delayed effects. Moreover, dry conditions have been found to increase Dengue risk in recent years. However, the impact of these factors on vector (mosquito) activity remains unclear. This study assessed the effect of human mobility and rainfall on the Dengue mosquito. The Gravitrap indices were used to characterize local mosquito (*Aedes Albopictus*) abundance and extensiveness conditions. We used established Gravitrap indices to characterize mosquito abundance and extensiveness in Hong Kong. We found that i) the decrease in residential mobility might increase mosquito abundance and extensiveness; and ii) low rainfall (<50 mm) was associated with a higher risk of mosquito abundance after 4.5 months. Additionally, heavy rainfall was associated with increased mosquito activity risk.The future mosquito activity risk is expected to increase because of the relaxation of social distancing measures after the COVID-19 pandemic along with climate change. The results suggest that non-linear delayed effects of meteorological factors together with human mobility change can be used for the Dengue mosquito forecast. Social distancing may be a way to reduce the risk of *Aedes albopictus*.

## INTRODUCTION

Dengue fever (DF) is one of the most widespread mosquito-borne diseases, with an estimated 390 million infections each year (1). *Aedes aegypti* and *Aedes albopictus* are two Dengue vectors. Global warming can facilitate their spread, leading to increasing risk in certain previously non-endemic subtropics, including southern China (2). In addition, according to a recent study, the risk of DF incidence can also be largely influenced by social distancing measures and human movement behaviours (3). Hong Kong, a metropolitan in southern China, faces an increased risk of Dengue outbreaks (4). Understanding how human mobility together with weather conditions affects *Aedes* mosquito abundance in Hong Kong helps make an early assessment of Dengue risk and decision-making in vector control.

In response to the increasing risk of DF (4), Hong Kong has established a Gravidtrap system for vector surveillance since 2020 to replace the Ovitrap. The Gravitrap was developed to capture female *Aedes Albopictus* (5). In addition to the extensiveness (i.e. the distribution) of *Aedes Albopictus*, which could be measured by both traps, the new trap also monitored the abundance (i.e. the number) of mosquitoes.

During 2020 and 2022, social distancing was regularly introduced to reduce the spread of COVID-19. In 2022, strict social distancing was introduced during the COVID-19 Omicron wave. The strict social distancing might affect the likelihood of mosquitoes biting humans, especially *Aedes albopictus*, which tends to stay outside (6). Therefore, incorporating human mobility into mosquito prediction modelling is essential, especially when social distancing is implemented.

While a warm condition has a known impact on mosquito development, breeding, survival, etc., (7) the effects of rainfall are diverse, with both heavier and lower rainfall being associated with increased Dengue risk. Rainfall was an important Dengue risk factor (1,8), but recent studies found that drought conditions were also associated with a higher risk of Dengue incidence at long lead times of up to 5 months (9,10). Similarly, springtime rainfall, a few months before seasonal Dengue outbreaks, appeared to be negatively associated with annual Dengue incidence in Taiwan and Hong Kong (4,11). These previous studies suggested a varied relationship between hydroclimatic factors and Dengue incidence with delay effects (4,9–11). However, whether these delay effects occurred through influencing mosquito population dynamics remains largely unknown.

Our study aimed to assess the influence of human mobility on the abundance and extensiveness of *Aedes albopictus*, taking account of the nonlinear lagged effects of total rainfall and mean temperature (Figure 1). The results provided important insights for Dengue risk prediction and control.

**Figure 1.**
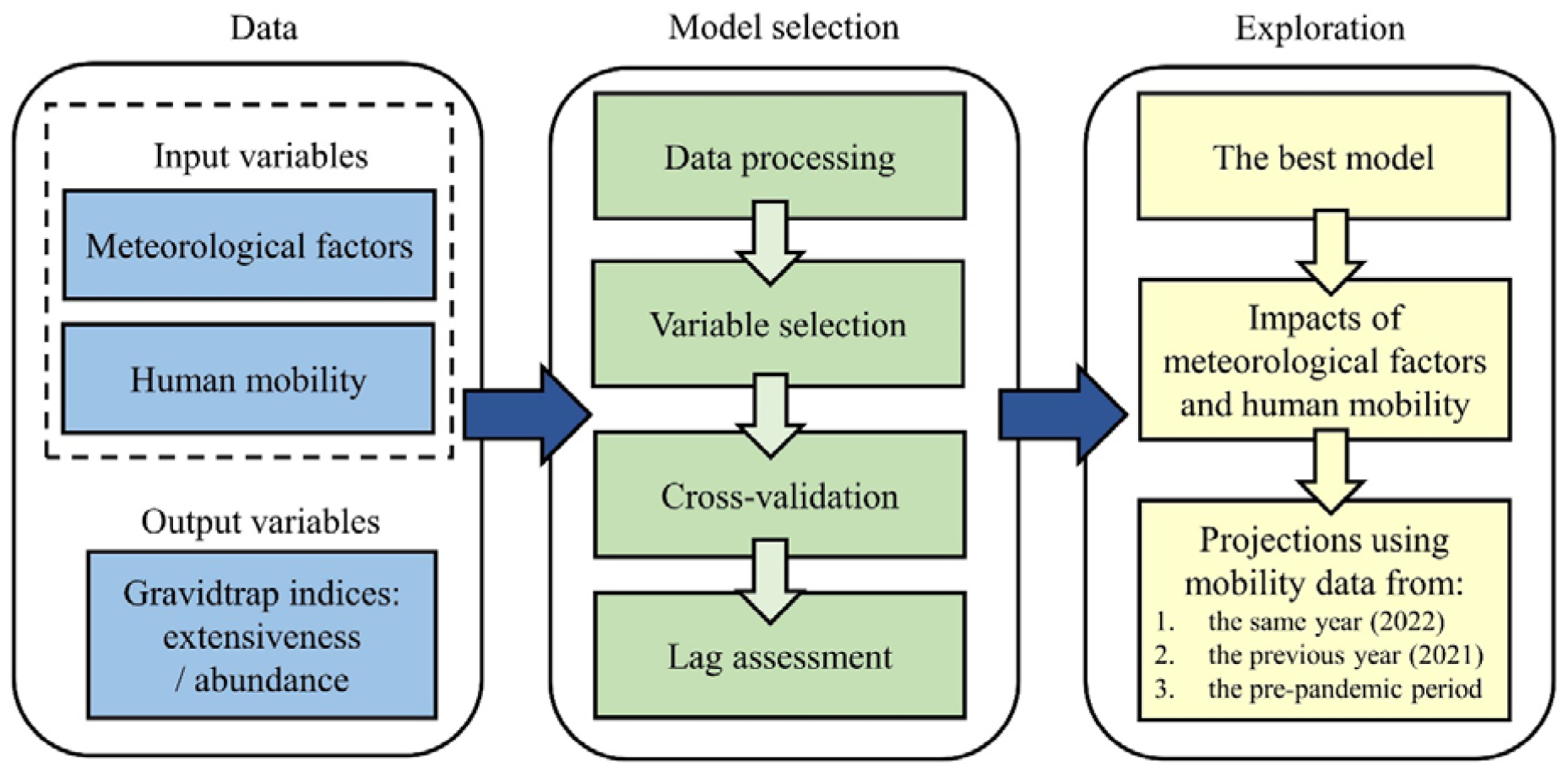
Schematic flow for exploration of different factors in the mosquito extensiveness and abundance predictions. WAIC was used for variable selection. The best model was determined after the results were cross-validated and compared with different lagged periods.

## METHODS

### Meteorological data

Meteorological data in Hong Kong were collected based on the weather stations in three regions from the Hong Kong Observatory (12), including daily total rainfall and daily mean temperature. We collected the data from April 2020 to August 2022 and divided it into three areas (13): Hong Kong Island and Kowloon (HKK), New Territories East (NTE), and New Territories West (NTW) (Table S1). Monthly total rainfall and monthly mean temperature (i.e. obtained by averaging the daily mean temperatures) were calculated from these daily measurements (Table S2). The meteorological factors for each area are the average value of selected weather stations within that area (Supplementary Methods).

### Human mobility data

Human mobility data for Hong Kong were collected from Google to represent the human behavioural changes in response to the COVID-19 pandemic and social distance measures during the study period (14). We used three categories of places in human mobility data, including residential, parks and workplaces. All indices were computed relative to a baseline day. The baseline day is the median value from the 5 weeks Jan 3–Feb 6, 2020. The monthly human mobility index was calculated for model prediction (see Supplementary Methods).

### Mosquito activity data

Mosquito activity data were provided by the Food and Environmental Hygiene Department (15). Two indices were measured by the mosquito Gravitrap surveillance: The Area Density Index (ADI) defined as the number of mosquitoes captured in the traps (used to represent the abundance of *Aedes albopictus*); and the Area Gravidtrap Index (AGI), defined as the proportion of Gravidtraps that are found to have positive results in a specific area (used to represent the extensiveness of *Aedes albopictus*).

We calculated the abundance and extensiveness for each area during each month *t*. First, the index for each area (i.e. HKK, NTE, and NTW) was calculated as the average index for all Gravidtrap sites in the region.

The average monthly mosquito extensiveness in area *A* is defined as:

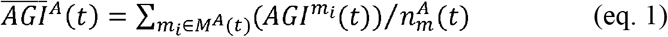

where 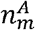 represents the total number of mosquito monitoring sites in area *A*. 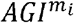 represents the AGI in the surveyed area *m*_*i*_. *M*^*A*^ = {*m*_1_, *m*_2_,…, *m*_*i*_} is a collection of sites (surveyed areas), in which *m*_*i*_ represents each site in the area.

The monthly abundance *N*^*A*^(*t*) (per 1,000 traps) in area *A* is defined as:

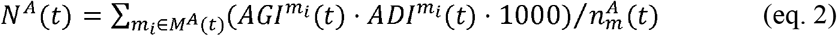

where 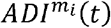 represents the mean ADI at the site *m*_*i*_ in month *t*.

### Model development

We developed prediction models for mosquito abundance and extensiveness based on distributed-lagged non-linear models (DLNM) (16). For mosquito abundance prediction, we assumed the mosquito number per 1000 traps follows the negative binomial distribution and selected the log function as the link function. Let *λ*_*t*_ be the mosquito number per 1000 traps in the month *t*, such as:

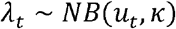

where *u*_*t*_ is distribution mean of *λ*_*t*_ at month *t* and *κ* is the overdispersion parameter in the negative binomial distribution. *λ*_*t*_ in area, *A* can be measured by monthly mosquito abundance *N*^*A*^(*t*) (see eq. 2). Then, the regression model to predict mosquito abundance was:

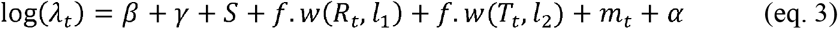

where *f·w*(*R*_*t*_, *l*_1_) and *f·w*(*T*_*t*_, *l*_2_) represent the nonlinear exposure-lag functions of total rainfall *R*_*t*_ from 0 to *l*_1_ months and mean temperature *T*_*t*_ from 0 to *l*_2_ months in *t*^*th*^ month, respectively; *S* is the area random effect; *γ* is the monthly random effect; *β* is the yearly random effect; *α* is the intercept; and *m*_*t*_ is the human mobility index in one category (e.g. residential areas, workplaces, or parks) at *t*^*th*^ month.

In the mosquito extensiveness prediction, we assumed the number of positive traps follows the binomial distribution and selected the logit function as the link function. Let *Y*_*t*_ be the number of positive traps in the *t*^*th*^ month, following the binomial distribution with the total number of the traps (*n*_*t*_), and the probability of a positive trap (*p*_*t*_) in the *t*^*th*^ month, such as:

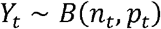

where *p*_*t*_ at each surveyed area *A* can be measured by 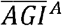 (see eq. 1). *n*_*t*_ can be calculated as the product of the number of surveyed sites and the average number of traps per site. In Hong Kong, an average of 55 Gravitrap were placed in each selected site. Similarly, the regression model to predict mosquito extensiveness was:

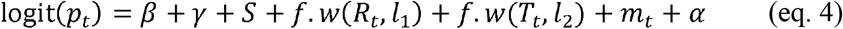

### Model selection criteria

To determine the best model (best-prediction model) among different combinations of predictors, in a two-stage selection approach, the Watanabe-Akaike information criterion (WAIC) was used in the first stage to select different sets of variables (candidate models) (17). Besides the yearly, monthly, and area-specific random effects, variables include the lag effects of total rainfall and mean temperature and three human mobility indices (see Table S3) In the second stage, leave-one-out cross-validation (LOOCV) based on mean square error (MSE) was performed for the candidate models to compare the model’s predictions to the observed data.. In LOOCV, the data set is divided into k parts (i.e. the total number of months), where k-1 parts are used as the training set and the remaining parts are used as the validation set. The procedure was repeated until every part (i.e. month) had been used for validation.

### Comparison between mosquito abundance and extensiveness

For comparing the predictive performance among mosquito abundance and extensiveness, we used two standardized mosquito indices to unify the scales of different indices: the standardized abundance index (SAI) and the standardized extensiveness index (SEI). The SAI was:

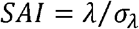

where *λ* and *σ*_*λ*_ represent mosquito abundance and standard deviation of mosquito abundance, respectively. Similarly, the SEI was:

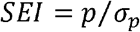

where *p* and *σ*_*p*_ represent mosquito extensiveness and standard deviation of mosquito extensiveness, respectively.

## RESULTS

### Data analysis

In Hong Kong, mosquito abundance exhibited strong seasonal patterns, growing in the spring (March-May), peaking in the early summer (June or July), and remaining nearly at 0 in the winter (December-February) (Figure 2A). Among three predefined regions, NTE (a northeastern region) recorded higher mosquito abundance (i.e. the number of *Aedes albopictus*) than others. Mosquito extensiveness (i.e. the distribution of *Aedes albopictus*) had a similar pattern of variation to abundance (Figure S1 and Figure S2). Two mosquito standardized indices were proposed for mosquito abundance and extensiveness prediction (Figure S3). in Hong Kong from April 2020 to August 2022. During the study period, human mobility generally fluctuated at the beginning but showed rapid declines in parks and workplaces and a sharp increase in residential since January 2022. This rapid change reflected the behavioural changes and social distancing measures introduced during the COVID-19 Omicron wave (Figure 2B).

**Figure 2.**
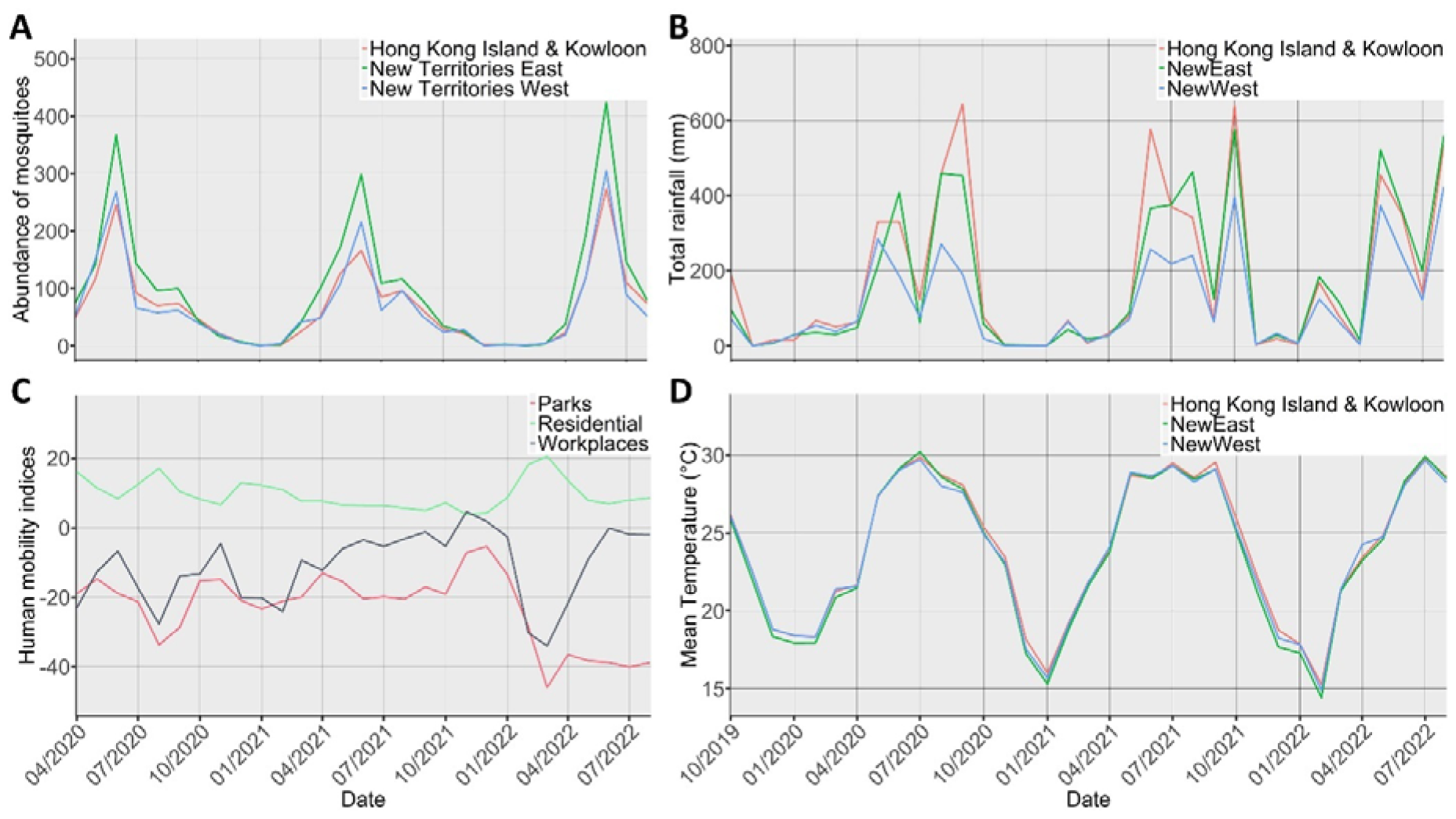
The monthly change in mosquito abundance and its predictors in Hong Kong. (A) Mosquito abundance; (B) Total rainfall; (C) Human mobility; (D) Mean temperature. In A, B, and D, red refers to Hong Kong Island and Kowloon (HKK); blue refers to New Territories East (NTE); and green refers to New Territories West (NTW).

The monthly total rainfall typically exceeded 300mm during summer and early autumn (June-October) and was mostly less than that during winter and early spring (November-April) but varied substantially between different regions and years. For example, in 2020–2021, HKK (a southern region) had the most total rainfall, while NTW (a northwestern region) had the least (Figure 2C). In 2022, all the regions experienced higher levels of total rainfall in February than the previous year. The monthly mean temperature among these three regions was similar, with the highest at about 30°C around July and the lowest at about 15°C in January or February (Figure 2D).

### Model selection of mosquito abundance and extensiveness

After initial variable selection, a baseline model for mosquito abundance prediction (Model A; see Table 1) was obtained. The candidate models in mosquito abundance prediction (see eq. 3) include Model A, Model A-M_p_, Model A-M_w_, and Model A-M_r_. After incorporating human mobility in residential, the best model for abundance (Model A-M_r_) was obtained using LOOCV. The best model for extensiveness (Model E-M_r_) also contained the same set of variables.

**Table 1.** Comparison of candidate models for mosquito abundance. The top six rows represent the results of variable selection to obtain the best model. The bottom six rows represent the results of sensitivity analysis to ensure the best time lag of total rainfall and mean temperature. Model A comprised the total rainfall with lags from 0 to 6 months, mean temperature with lags from 0 to 2 months, and random effects for years, months, and regions were used as a baseline model. Model A-M_p_, Model A-M_w_, and Model A-M_r_ were models incorporating the human mobility index in parks, workplaces, and residential based on Model A, respectively.

| Model | Model formula | WAIC | MSE in LOOCV |
| --- | --- | --- | --- |
| Model A1 | $\beta + \gamma + S + \alpha$ | 741.12 | — |
| Model A2 | $\beta + \gamma + S + f.w(R_t, 6) + \alpha$ | 727.31 | — |
| Model A | $\beta + \gamma + S + f.w(R_t, 6) + f.w(T_t, 2) + \alpha$ | 681.60 | 0.63 |
| Model A-M<sub>p</sub> | $\beta + \gamma + S + f.w(R_t, 6) + f.w(T_t, 2) + m_p + \alpha$ | 677.92 | 0.65 |
| Model A-M<sub>w</sub> | $\beta + \gamma + S + f.w(R_t, 6) + f.w(T_t, 2) + m_w + \alpha$ | 658.89 | 0.46 |
| Model A-M<sub>r</sub> | $\beta + \gamma + S + f.w(R_t, 6) + f.w(T_t, 2) + m_r + \alpha$ | **651.87** | **0.37** |
| Model A-M<sub>r</sub>1 | $\beta + \gamma + S + f.w(R_t, 5) + f.w(T_t, 2) + m_r + \alpha$ | 666.57 | — |
| Model A-M<sub>r</sub>2 | $\beta + \gamma + S + f.w(R_t, 4) + f.w(T_t, 2) + m_r + \alpha$ | 668.37 | — |
| Model A-M<sub>r</sub>3 | $\beta + \gamma + S + f.w(R_t, 3) + f.w(T_t, 2) + m_r + \alpha$ | 664.26 | — |
| Model A-M<sub>r</sub>4 | $\beta + \gamma + S + f.w(R_t, 2) + f.w(T_t, 2) + m_r + \alpha$ | 671.56 | — |
| Model A-M<sub>r</sub>5 | $\beta + \gamma + S + f.w(R_t, 1) + f.w(T_t, 2) + m_r + \alpha$ | 669.79 | — |
| Model A-M<sub>r</sub>6 | $\beta + \gamma + S + f.w(R_t, 6) + f.w(T_t, 1) + m_r + \alpha$ | 653.22 | — |

### Prediction of mosquito activity using weather and human mobility

Overall, both models A-M_r_ and E-M_r_ showed similar predictive performances (Table 1 and Table S4), The model fit for mosquito abundance appeared to be slightly better than that for mosquito extensiveness as more observed data points in the year 2022 were within 95% confidence interval (CI) of the predicted values for mosquito abundance (Figure 3A and Figure S4A). It might be because mosquito abundance is more weather-related while mosquito extensiveness is more related to the presence of potential breeding places or the placement of the traps (Table 1 and Table S4).

**Figure 3.**
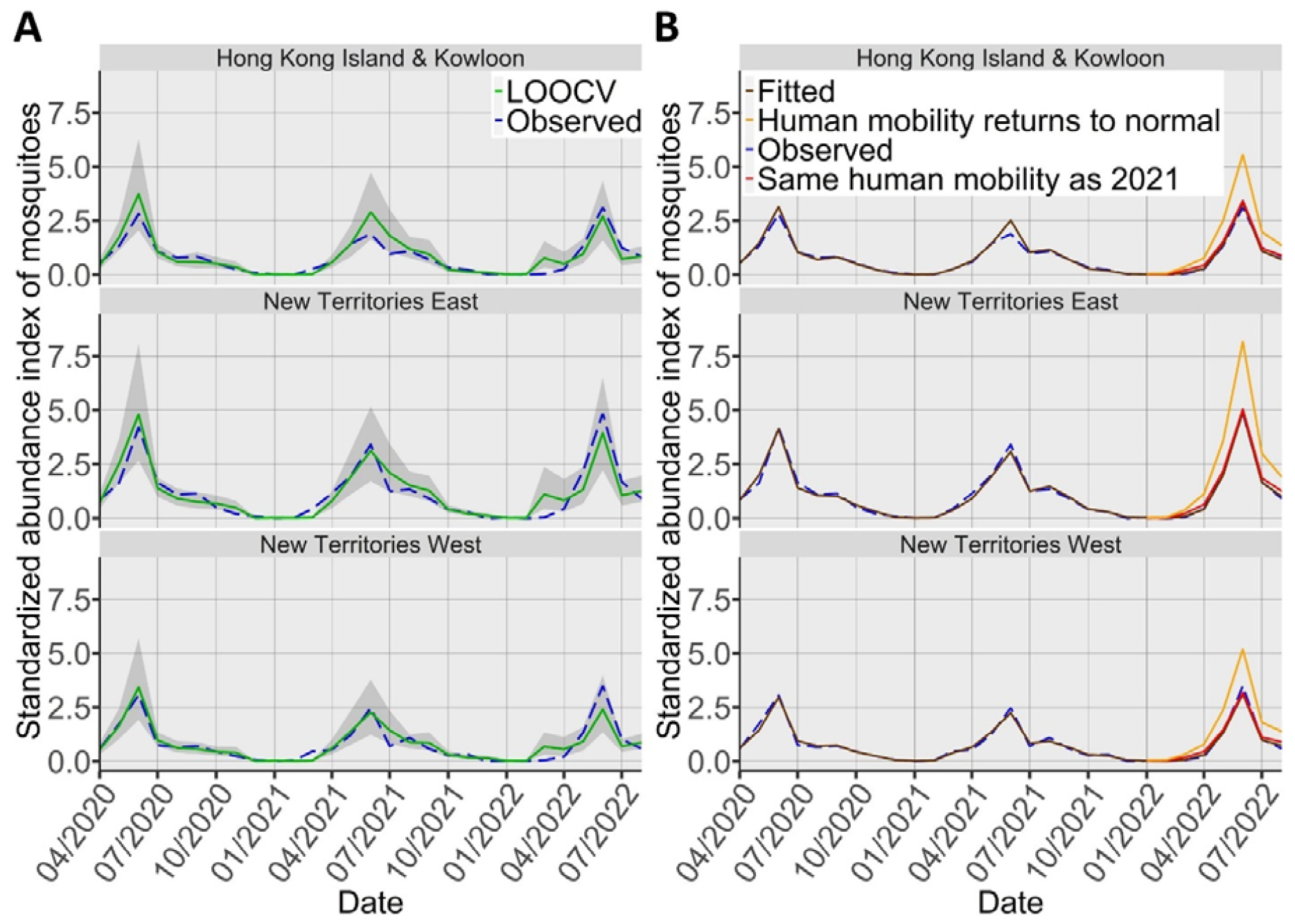
Comparison of observed and predicted results using the best model for mosquito abundance. (A) Predicted results of mosquito abundance using leave-one-out cross-validation (LOOCV) with Model A-M_r_. The grey shaded area represents the 95% confidence interval. The blue dashed line represents observed data and the green solid line is the leave-one-out cross-validation result. (B) Projected results of mosquito abundance in the year 2022 under different scenarios in human mobility (residential category): human mobility returns to the COVID-19 pre-pandemic period (orange) and the same human mobility as year 2021 (red). The blue dashed line represents observed data and the brown solid line represents fitted data.

The model estimated that residential mobility change has a negative effect on the mosquito abundance prediction (correlation coefficient = -0.075 (95%CI, -0.118--0.033); see Table S5). On the other hand, park mobility and workplace mobility had a positive effect on mosquito abundance. The effects of the human mobility indices on mosquito extensiveness were consistent with mosquito abundance.

### Effects of weather conditions

Higher relative risks in mosquito abundance were observed in the conditions of extremely low (<50 mm) or heavy rainfall (>500 mm) (Figure 4A). Compared with the reference (300 mm), a reduction in total rainfall was associated with a higher relative risk (RR) after about 4.5 months, reaching a maximum of 1.73 (95%CI, 1.19-2.51). However, heavy rainfall conditions (>500 mm) were associated with a higher RR within 3 months, reaching a maximum of 1.31 (95%CI, 0.99-1.73). When lagged effects were accumulated, the maximum RR occurred at no rainfall (Figure 4B). The accumulated RR decreased with total rainfall up to about 500 mm but increased again thereafter.

**Figure 4.**
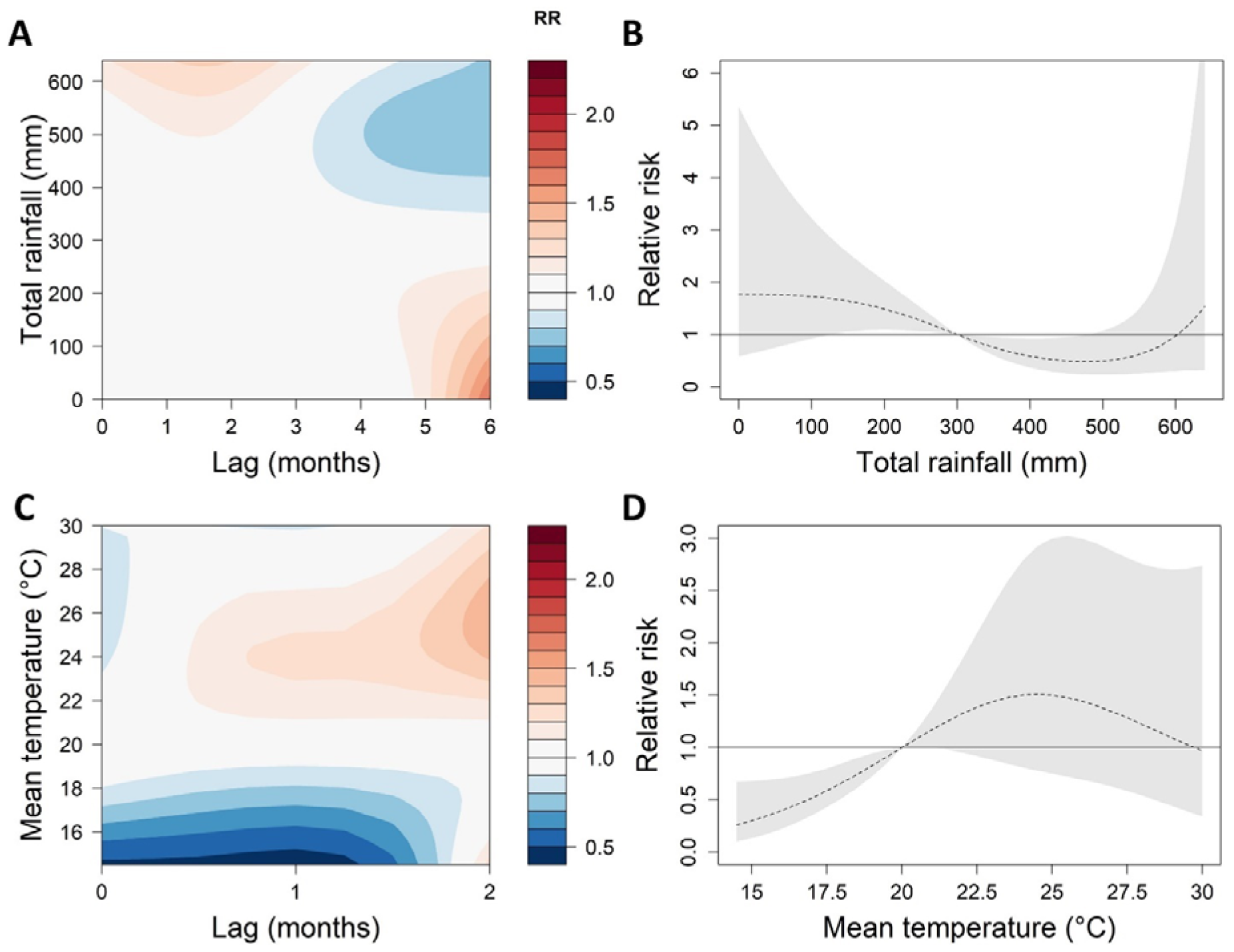
Effects of total rainfall and mean temperature on mosquito abundance using the best model. (A) Relative risk (RR) by total rainfall and lag months; (B) Cumulative RR for total rainfall; (C) RR by mean temperature and lag months; (D) Cumulative RR for mean temperature. The reference of total rainfall and mean temperature are 300mm and 20°C, respectively. The deeper the shade of red, the greater the increase in RR compared with the reference. The deeper the shade of blue, the greater the decrease in RR compared with the reference. The black dashed line represents the cumulative exposure-response association. The grey shaded area represents the 95% confidence interval.

Compared with the reference mean temperature (20 °C), the warm conditions (21-30 °C) were associated with an increased risk of mosquito abundance after about half a month, leading to a maximum RR of 1.47 (95%CI, 0.94-2.32) at 25.5 °C (Figure 4C). When lagged effects were accumulated, the RR increased with mean temperatures from 15 °C to 26 °C but decreased at warmer conditions (>26 °C) (Figure 4D). The associations between total rainfall and mean temperature with mosquito extensiveness were similar to that of mosquito abundance (Figure S5).

### Model projections for human mobility change

The model projected the mosquito abundance in two alternative scenarios for the year 2022: 1) same residential mobility as before the COVID-19 Omicron wave, indicating returning to a normal state, and 2) same residential mobility as year 2021, indicating a weak social distancing. In the first scenario when human mobility returned to normal (before the COVID-19 pandemic), cumulative mosquito abundance in all areas significantly increased by an average of 80.49% compared to the actual situation. The cumulative mosquito abundance in the HKK is increasing the most, at 83.41%. In the second scenario, cumulative mosquito abundance in all areas was only slightly higher than the actual situation by an average of 10.96% (Figure 3B). The projection results of mosquito extensiveness were similar to those of mosquito abundance (Figure S4B).

### Sensitivity analysis

Prediction models in mosquito abundance and extensiveness were sensitive in the variable selection of the lag of total rainfall, the lag of mean temperature, and the human mobility indices (Table 1 and Table S4). Model A, the WAIC decreased from 681.60 to 651.88 after adding the residential mobility index. When the total rainfall lag reduced from 6 months to other lengths, WAIC increased. Similarly, When the mean temperature lag reduced from 2 months to 1 month, WAIC increased. The durations of lags in total rainfall and mean temperature that produced the lowest WAIC for mosquito abundance and extensiveness were selected.

## DISCUSSION

### Influence of human mobility on mosquito activity

Understanding the association between human mobility and mosquito activity helps in forecasting the risk of DF transmission and developing effective strategies for its prevention. Our results showed that residential mobility was negatively associated with mosquito abundance, while mobility indices in parks and workplaces showed positive associations. The results can be explained by that the increase in outdoor activities provides more chances to feed meals (blood) to *Aedes albopictus* (6). The model projected that if human mobility (i.e. residential) returned to normal in 2022, the risk of mosquito activity would increase significantly (80% more) during the peak. In addition to border reopening (18), a possible increase in mosquito abundance, extensiveness, and DF incidence might appear due to more outdoor human activity.

Recently, Brady et al. also reported that Dengue incidence was associated with certain Google human mobility indices (e.g. workplace mobility and park mobility) during the COVID-19 period (3). The difference between their modelling results with our study may be explained by some factors. The impact of human mobility may be different in mosquito activity and Dengue incidence. Moreover, an increase in residential mobility may have different impacts on mosquitoes between *Aedes Albopictus* and *Aedes Aegypti*, which are both the main vectors to transmit DF. However, *Aedes Albopictus* was the only mosquito vector found in Hong Kong. Those differences suggest that prevention of DF through vector control should take into account the impact of human mobility on mosquito and Dengue incidence simultaneously.

### Impact of total rainfall lags and mean temperature on mosquito activity

Rainfall appeared to influence mosquito activity by different mechanisms. Both dry and wet conditions were found to be associated with an increased risk of Dengue infection in China (19). We found that heavy rainfall conditions (>500 mm) within 3 months were associated with a higher risk of *Aedes* mosquito activity. On the other hand, low rainfall (<50 mm) was associated with a higher risk with a longer lag (Figure 4A). A possible explanation is that early low rainfall (around late winter or springtime) helps to maintain the number of mosquito eggs or larval sites without the flushing effect (20). *Aedes albopictus* survives the winter at its egg stage. When more rain comes in a warmer environment after a period of dry conditions, there will be a large number of eggs hatching simultaneously leading to a bloom in adult mosquitoes. This scenario can be particularly important when the mosquito begins to grow, corresponding to the period from springtime to the pre-rainy season before the monsoon in many subtropical regions in Asia (21). The results suggest that, in southern China or its neighbourhoods, some extreme hydroclimatic events, such as delayed monsoon or drought conditions, during springtime might increase the Dengue risk in summertime. Severe drought conditions have been observed in spring in 2018 in Hong Kong, and 2015 and 2023 in Taiwan, followed by significant outbreaks (11,22,23).

In our findings, the mean temperature was positively associated with mosquito activity until exceeding a threshold. We observed an increased mosquito activity risk after about a half-month, similar to the time that *Aedes albopictus* may use to develop from eggs to adults. A maximum level occurred about two months later when the mean temperature was between 25 and 26°C (Figure 4C). Our findings in Hong Kong were consistent with the previous studies using Dengue incidence (9).

### Implications

The spread of vector-borne diseases such as DF, Malaria, and Zika diseases can have serious consequences for human health and even lead to large-scale deaths. Residential mobility indicates the change in the time people spend at home, as the outcome of social distancing measures during the COVID-19 pandemic period. Therefore, our results suggest that social distancing measures may be an important intervention to reduce mosquito abundance. Furthermore, our results indicate that knowing the impact of hydroclimatic events on these diseases can provide important risk projections for the future. In vector control of South-East Asia, the Global Vector Control Response prioritized enhancing vector surveillance, forecasting, and monitoring effects of different factors (24). Extreme weather or hydroclimatic events, such as heavy rains or droughts are happening more frequently in the world (25). They might have critical influences on the dynamics of the mosquito population depending on the time of their occurrences (26). Incorporating extreme hydroclimatic events (such as drought conditions) together with human mobility patterns helps to forecast Dengue risk and inform public health decisions in vector control for its prevention.

### Limitations and future works

Several limitations exist within this study. The mosquito activity may also be influenced by other human-induced factors, such as land use type and urbanization process (27,28). Control measures may have been taken specifically around those Gravidtrap sites. But during the zero-COVID period in Hong Kong, the impact of these mosquito control measures was expected to be smaller. Furthermore, extreme weather events can affect a variety of climatic factors, some of which might be important factors for mosquito activity, such as typhoons (29). We chose two important weather factors according to the previous studies (1,8), instead of considering all of them. Recent studies have assessed the impact of climate change on Dengue risk (19,30). Incorporating the insights from our findings can provide a better understanding of how extreme weather conditions and human mobility might influence the Dengue vector’s abundance and control.

## Conclusion

As the COVID-19 pandemic ends and borders reopen, the risk of DF is expected to be higher in many parts of the world. The study found that social distancing measures were associated with reduced mosquito abundance and extensiveness. Furthermore, low rainfall was associated with a higher risk of mosquito activity (abundance and extensiveness) with the 4.5 months lag (Figure 4A), which was able to explain the recent findings of the delayed effects of weather conditions on Dengue incidence (9,10). The results suggested that the highest Dengue risk after drought conditions was likely to be affected by the spread of Dengue mosquitoes.

## Data Availability

All data produced are available online at https://github.com/YufanZheng/Mosquito-Association-Modeling.

## Contributors

YZ, KY and HY designed the study. YZ and KY developed the methods, performed the research, and accessed and verified the data. YZ and KY wrote the initial draft of this manuscript. YZ and HY contributed to the interpretation of the results. YZ, HY, and EW helped revise the manuscript. All authors had full access to all the data in the study and had final responsibility for the decision to submit for publication.

## Acknowledgements

We thank Ming Wai Lee from the Food and Environmental Hygiene Department, the Govern ment of HKSAR.

