## Supplementary for "Association of Human Mobility and Weather Conditions with Dengue Mosquito Abundance during the COVID-19 Pandemic in Hong Kong"

**SUPPLEMENTARY MATERIAL**

**SUPPLEMENTARY METHODS**

**Meteorological data**

In month $t$, there are $n_{w}^{A}(t)$ weather stations in area $A$. The monthly value of weather factors is the average of the daily value in each month. We assumed that $W^{A}\left( t \right)=\{w_{1},w_{2},\ldots,w_{j}\}$ is a collection of stations in area $A$, where $w_{j}$ represents one station in this area. Then, we calculated the mean temperature and total rainfall in each area.

The monthly mean temperature $T^{A}\left( t \right)$ in area $A$:

$$T^{A}(t)=\frac{\sum_{w_{j}\in W^{A}(t)} T^{w_{j}}(t)}{n_{w}^{A}(t)}$$

Where and $T^{w_{j}}\left( t \right)$ represents the mean temperature in the station $w_{j}$ in month $t$.

The monthly total rainfall $R^{A}\left( t \right)$ in area $A$:

$$R^{A}(t)=\frac{\sum_{w_{j}\in W^{A}(t)} R^{w_{j}}(t)}{n_{w}^{A}(t)}$$

Where $R^{w_{j}}\left( t \right)$represents the total rainfall in the station $w_{j}$ in month $t$.

**Human mobility data**

We assumed that $H^{D}\left( t \right)=\{h_{1}(t),h_{2}(t),\ldots,h_{N}(t)\}$ is a collection of daily human mobility in month $t$ in Hong Kong, where $N_{t}$ is days of month $t$ and $D$ represents the category of human mobility. Then, the monthly human mobility index $m_{D}\left( t \right)$ in $D$ category in month $t$:

$$m_{D}\left( t \right)=\frac{\sum_{h_{i}(t)\in H^{D}\left( t \right)} h_{i}(t)}{N_{t}}$$

**SUPPLEMENTARY RESULTS**

**Prediction results of mosquito extensiveness**

In the first stage of the model selection, the candidate models in mosquito extensiveness prediction include Model E, Model E-M_p_, Model E-M_w_, and Model E-M_r_. Model E comprised the total rainfall with lags from 0 to 6 months, mean temperature with lags from 0 to 2 months, and random effects was used as a baseline model. Model E-M_p_, Model E-M_w_, and Model E-M_r_ were models incorporating the human mobility index in parks, workplaces, and residential based on Model E, respectively. After incorporating human mobility in residential, the best model for extensiveness (Model E-M_r_) was obtained using LOOCV (Figure S4A and Table S4). In the Model E-M_r_, the coefficient of the residential is -0.1032, which indicates the residential mobility change has a negative effect on the mosquito extensiveness prediction.

The model projected the mosquito extensiveness in two alternative scenarios for the year 2022: 1) same mobility as before the COVID-19 pandemic, indicating returning to the normal state, and 2) same mobility as year 2021, indicating a weak social distancing. In the first scenario, cumulative mosquito extensiveness increased by an average of 105.70% in the three areas (Figure S4B). The cumulative mosquito extensiveness in New Territories West was increasing the most, at 114.44%. In the second scenario, cumulative mosquito extensiveness slightly increased in the three areas (Figure S4B).

In the sensitivity analysis for meteorological lag variables, we compared the WAIC using different lag months of total rainfall and mean temperature in the best models (Table 1 and Table S4).

**SUPPLEMENTARY TABLES AND FIGURES**

**Table S1 The monitoring sites of mosquito activity data in three areas in Hong Kong**

| **Area** | **Site** |
| --- | --- |
| Hong Kong Island & Kowloon | Chai Wan West, Tin Hau, Shau Kei Wan & Sai Wan Ho, Wan Chai North, Happy Valley, Central, Sheung Wan and Sai Ying Pun, Sai Wan, North Point, Aberdeen and Ap Lei Chau, Pokfulam, Deep Water Bay & Repulase Bay, Cheung Chau, Tung Chung, Tsim Sha Tsui, Mong Kok, Lai Chi Kok, Sham Shui Po East, Cheung Sha Wan, Kowloon City North, Hung Hom, Ho Man Tin, Lok Fu West, Kai Tak North, Wong Tai Sin Central, Diamond Hill, Ngau Chi Wan, Kwun Tong Central, Lam Tin, Kowloon Bay |
| New Territories East | Tseung Kwan O South, Sai Kung Town, Tseung Kwan, Ma On Shan, Lek Yuen, Yuen Chau Kok, Tai Wai, Wo Che, Tai Po, Fanling, Sheung Shui |
| New Territories West | Tin Shui Wai, Yuen Kong, Yuen Long Town, Tuen Mun (S), Tuen Mun (N), Tuen Mun West, So Kwun Wat, Tsuen Wan Town, Tseun Wan West, Ma Wan, Sheung Kwai Chung, Kwai Chung, Lai King, Tsing Yi, Tsing Yi South, Tsing Yi North |

**Table S2 The weather stations in three areas of Hong Kong**

| **Areas** | **Weather Stations (temperature)** | **Weather Stations (rainfall)** |
| --- | --- | --- |
| Hong Kong Island & Kowloon | King’s Park, Happy Valley, Wong Chuk Hang | Quarry Bay, Cape D'Aguilar, Happy Valley, King’s Park |
| New Territories East | Ta Kwu Ling, Sha Tin, Tai Mei Tuk | Ta Kwu Ling, Sha Tin, Tai Mei Tuk |
| New Territories West | New Tsing Yi Station, Sha Lo Wan, Cheung Chau, Tuen Mun Children and Juvenile Home, Wetland Park | Sha Lo Wan, Cheung Chau, Tuen Mun Children and Juvenile Home, Wetland Park |

**Table S3 The variable for the predictive model**

| **Variable** | **Description** |
| --- | --- |
| $\beta$ | The yearly random effect |
| $\gamma$ | The monthly random effect |
| $S$ | The area random effect |
| $f.w\left( R_{t},l_{1} \right)$ | The nonlinear exposure-lag function of total rainfall $R_{t}$ with a lag of 0 to $l_{1}$ months |
| $f.w\left( T_{t},l_{2} \right)$ | The nonlinear exposure-lag function of mean temperature $T_{t}$ with a lag of 0 to $l_{2}$ months |
| $m_{p}$ | The monthly human mobility index in the parks |
| $m_{r}$ | The monthly human mobility index in residential places |
| $m_{w}$ | The monthly human mobility index in workplaces |

**Table S4 Comparison of candidate models for mosquito extensiveness.**

The top six rows represent the results of variable selection to obtain the best model. The bottom six rows represent the results of sensitivity analysis to ensure the best time lag of total rainfall and mean temperature. Model E comprised the total rainfall with lags from 0 to 6 months, mean temperature with lags from 0 to 2 months, and random effects for years, months, and regions were used as a baseline model. Model E-M_p_, Model E-M_w_, and Model E-M_r_ were models incorporating the human mobility index in parks, workplaces, and residential based on Model A, respectively.

| **Model** | **Model formula** | **WAIC** | **MSE in LOOCV** |
| --- | --- | --- | --- |
| Model E1 | $\beta+\gamma+S+\alpha$ | 1267.99 | — |
| Model E2 | $\beta+\gamma+S+f.w(R_{t},6)+\alpha$ | 1068.55 | — |
| Model E | $\beta+\gamma+S+f.w\left( R_{t},6 \right)+f.w(T_{t},2)+\alpha$ | 845.49 | 0.28 |
| Model E-M_p_ | $\beta+\gamma+S+f.w\left( R_{t},6 \right)+f.w(T_{t},2)+m_{p}+\alpha$ | 839.92 | 0.27 |
| Model E-M_w_ | $\beta+\gamma+S+f.w\left( R_{t},6 \right)+f.w(T_{t},2)+m_{w}+\alpha$ | 804.25 | 0.19 |
| Model E-M_r_ | $\beta+\gamma+S+f.w\left( R_{t},6 \right)+f.w(T_{t},2)+m_{r}+\alpha$ | **781.56** | **0.13** |
| Model E-M_r_1 | $\beta+\gamma+S+f.w\left( R_{t},5 \right)+f.w(T_{t},2)+m_{r}+\alpha$ | 829.06 | — |
| Model E-M_r_2 | $\beta+\gamma+S+f.w\left( R_{t},4 \right)+f.w(T_{t},2)+m_{r}+\alpha$ | 830.10 | — |
| Model E-M_r_3 | $\beta+\gamma+S+f.w\left( R_{t},3 \right)+f.w(T_{t},2)+m_{r}+\alpha$ | 817.21 | — |
| Model E-M_r_4 | $\beta+\gamma+S+f.w\left( R_{t},2 \right)+f.w(T_{t},2)+m_{r}+\alpha$ | 855.03 | — |
| Model E-M_r_5 | $\beta+\gamma+S+f.w\left( R_{t},1 \right)+f.w(T_{t},2)+m_{r}+\alpha$ | 846.24 | — |
| Model E-M_r_6 | $\beta+\gamma+S+f.w\left( R_{t},6 \right)+f.w(T_{t},1)+m_{r}+\alpha$ | 837.81 | — |

**Table S5 Mobility coefficient in different prediction models**

| **Model** | **Variable** | **Coefficient (95% CI)** |
| --- | --- | --- |
| Model A-M_p_ | $m_{p}$ | 0.0122 (-0.0007, 0.0253) |
| Model A-M_r_ | $m_{r}$ | -0.0753 (-0.1182, -0.0332) |
| Model A-M_w_ | $m_{w}$ | 0.0305 (0.0119, 0.0496) |
| Model E-M_p_ | $m_{p}$ | 0.0170 (0.0091, 0.0249) |
| Model E-M_r_ | $m_{r}$ | -0.1032 (-0.1311, -0.0755) |
| Model E-M_w_ | $m_{w}$ | 0.0386 (0.0264, 0.0509) |

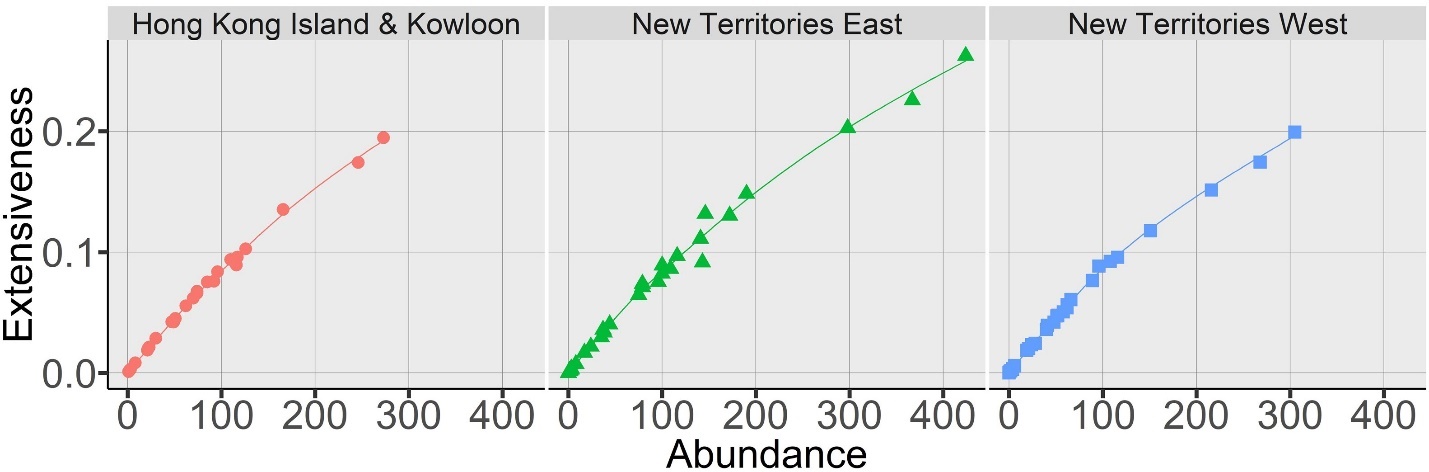
**Figure S1 The relationship between the extensiveness and abundance of mosquitoes in Hong Kong.** The dot is the value of a particular month in an area. Red refers to Hong Kong Island and Kowloon; blue refers to New Territories East; and green refers to New Territories West. The line is the curve-fitting line between extensiveness and abundance by using local weighted linear regression.

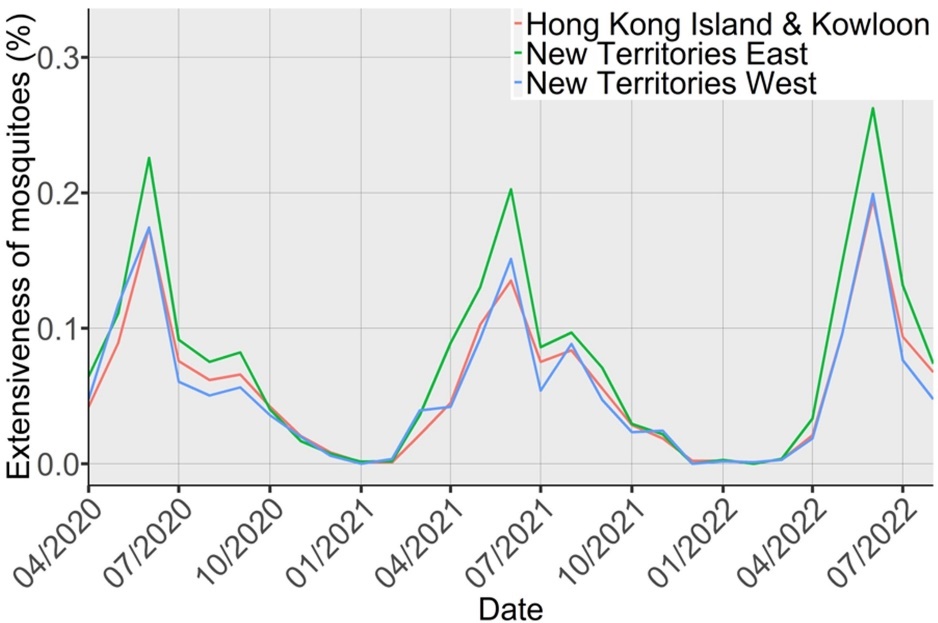

**Figure S2 The monthly change of mosquito extensiveness in Hong Kong.** Red refers to Hong Kong Island and Kowloon; blue refers to New Territories East; and green refers to New Territories West.

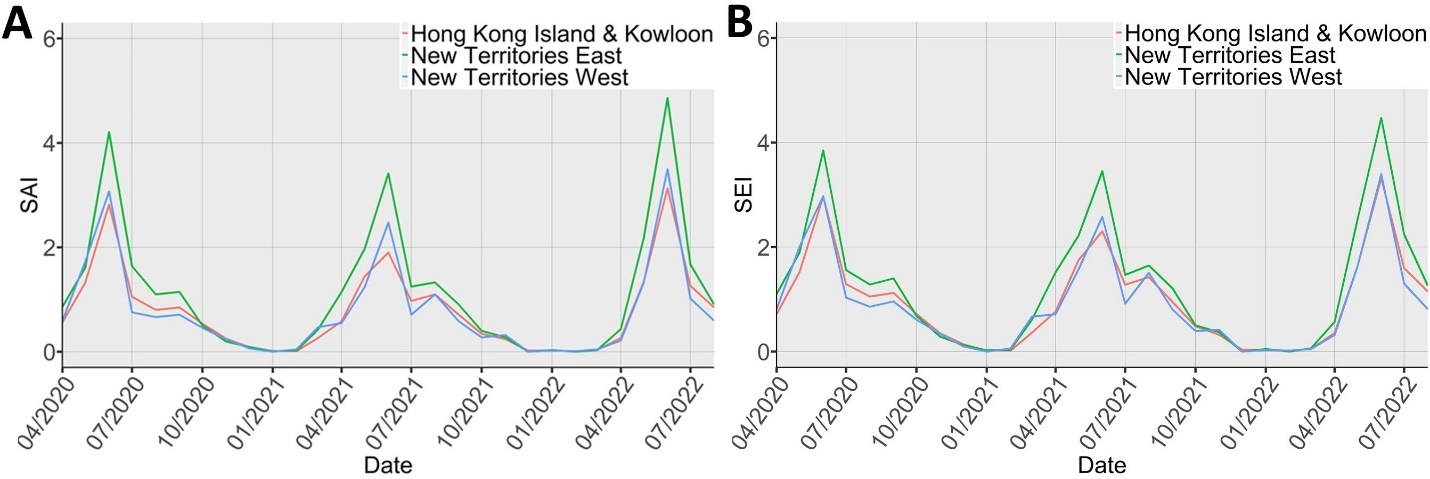
**Figure S3 Two standardized indices in three areas in Hong Kong.** (A) Standardized abundance index of mosquitoes (SAI); (B) Standardized abundance index of mosquitoes (SEI). Red refers to Hong Kong Island and Kowloon; blue refers to New Territories East; and green refers to New Territories West.

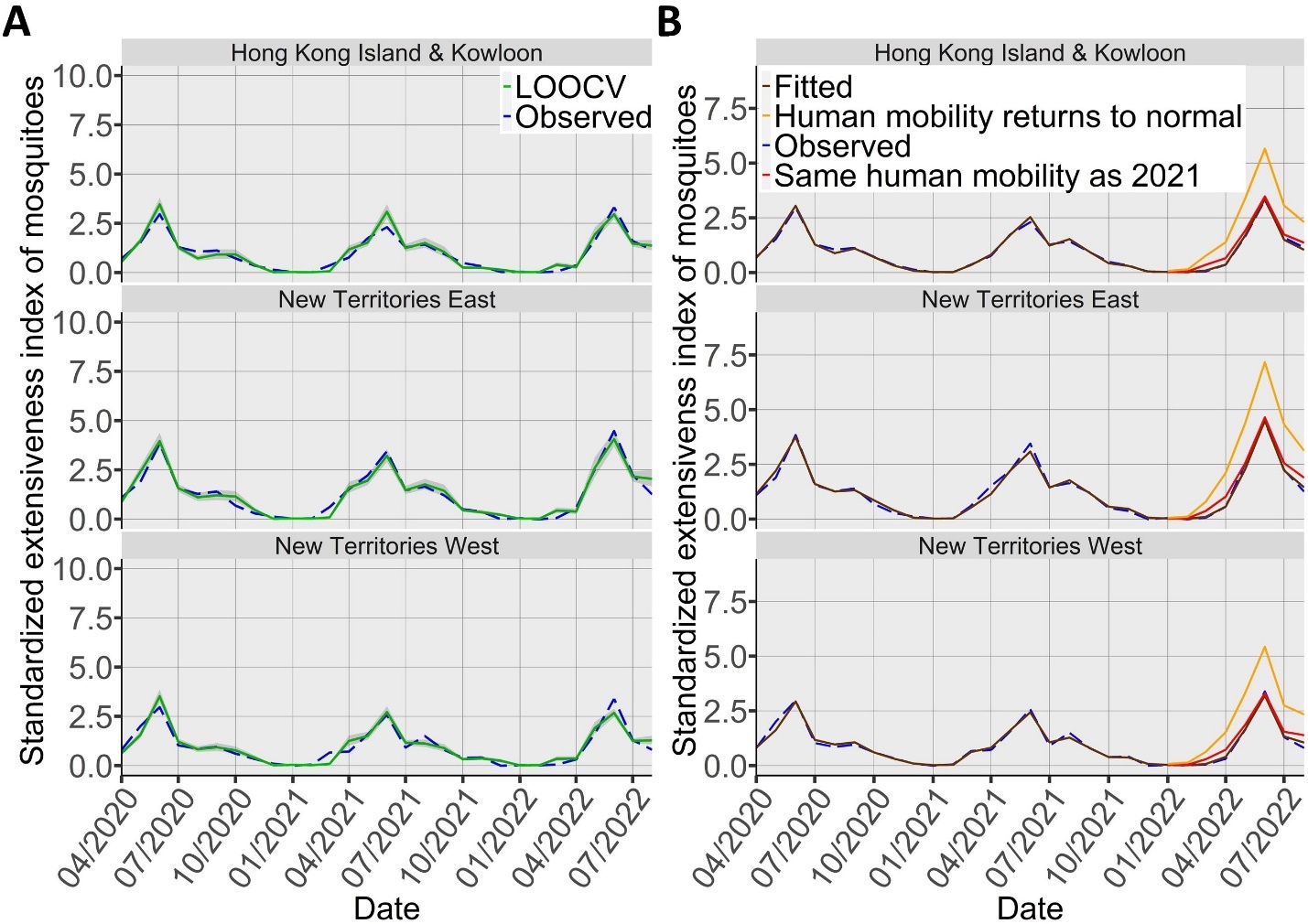

**Figure S4 Comparison of observed and predicted results using the best model for mosquito extensiveness.** (A) Predicted results of mosquito extensiveness using leave-one-out cross-validation (LOOCV) with Model E-Mr. The grey shaded area represents the 95% confidence interval. The blue dashed line represents observed data and the green solid line is the leave-one-out cross-validation result. (B) Projected results of mosquito extensiveness in the year 2022 under different scenarios in human mobility (residential category): human mobility returns to the COVID-19 pre-pandemic period (orange) and the same human mobility as the year 2021 (red). The blue dashed line represents observed data and the brown solid line represents fitted data.

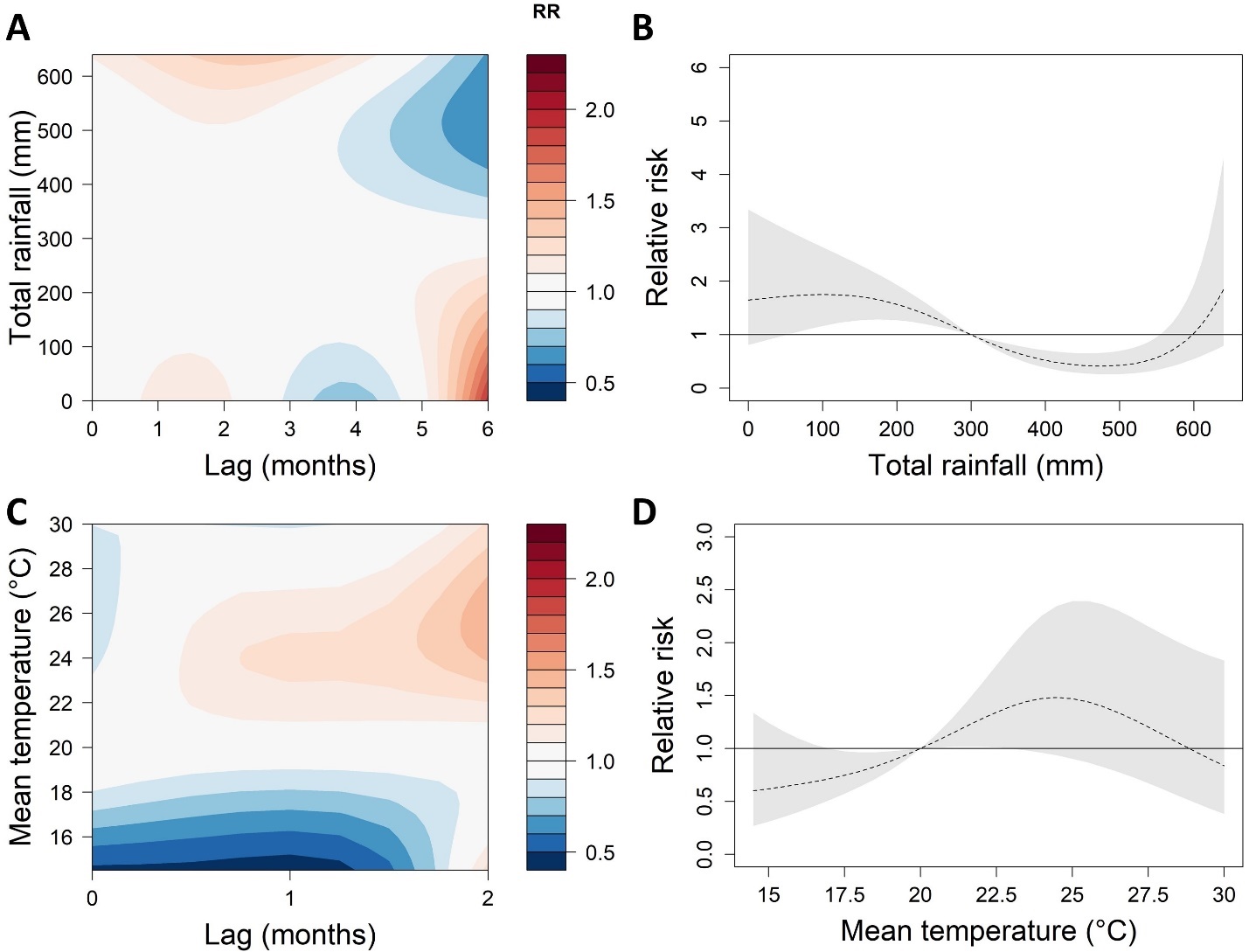
**Figure S5 Effects of total rainfall and mean temperature on mosquito extensiveness using the best model.** (A) Relative risk (RR) by total rainfall and lag months; (B) Cumulative RR for total rainfall; (C) RR by total rainfall and lag months; (D) Cumulative RR for mean temperature. The reference of total rainfall and mean temperature are 300mm and 20^◦^C, respectively. The deeper the shade of red, the greater the increase in RR compared with the reference. The deeper the shade of blue, the greater the decrease in RR compared with the reference. The black dashed line represents the cumulative exposure-response association. The grey shaded area represents the 95% confidence interval.
